# Assessing COVID-19 vaccination strategies in varied demographics using an individual-based model

**DOI:** 10.1101/2022.06.11.22276248

**Authors:** Noam Ben-Zuk, Yair Daon, Amit Sasson, Dror Ben-Adi, Amit Huppert, Daniel Nevo, Uri Obolski

## Abstract

**Background:** New variants of SARS-CoV-2 are constantly discovered. Administration of COVID-19 vaccines and booster doses, combined with applications of non-pharmaceutical interventions (NPIs), is often used to prevent outbreaks of emerging variants. Such outbreak dynamics are further complicated by the population’s behavior and demographic composition. Hence, realistic simulations are needed to estimate the efficiency of proposed vaccination strategies in conjunction with NPIs.

**Methods:** We developed an individual-based model of COVID-19 dynamics that considers age-dependent parameters such as contact matrices, probabilities of symptomatic and severe disease, and households’ age distribution. As a case study, we simulate outbreak dynamics under the demographic compositions of two Israeli cities with different household sizes and age distributions. We compare two vaccination strategies: vaccinate individuals in a currently prioritized age group, or dynamically prioritize neighborhoods with a high estimated reproductive number. Total infections and hospitalizations are used to compare the efficiency of the vaccination strategies under the two demographic structures, in conjunction with different NPIs.

**Results:** We demonstrate the effectiveness of vaccination strategies targeting highly infected localities and of NPIs actively detecting asymptomatic infections. We further show that there are different optimal vaccination strategies for each demographic composition of sub-populations, and that their application is superior to a uniformly applied strategy.

**Conclusion:** Our study emphasizes the importance of tailoring vaccination strategies to subpopulations’ infection rates and to the unique characteristics of their demographics (e.g., household size and age distributions). The presented simulation framework and our findings can help better design future responses against the following emerging variants.

## Introduction

In December 2019, a new virus, SARS-CoV-2, emerged in Wuhan, China. In January 2020, the World Health Organization (WHO) announced coronavirus disease 19 (COVID-19) a pandemic (1). Countries closed their borders and implemented harsh travel restrictions to try and slow down the spread of the virus (2). Nevertheless, the virus led to massive mortality, severe hospital loads, and a worldwide impact on the economy (1). At the beginning of the pandemic, a vaccine was not available. Hence, to relieve the burden on hospitals and limit the number of casualties governments worldwide applied non-pharmaceutical interventions (NPIs). These NPIs included home isolation of infected or suspected individuals, closure of workplaces, schools, kindergartens, and more (3).

The Food and Drug Administration (FDA) approved the first SARS-CoV-2 vaccine at the end of 2020, and a worldwide vaccination operation started. Since then, 4.6 billion people have been fully vaccinated and 11.8 billion doses have been administered worldwide (4). Countries chose different vaccination strategies, including rollout rates, and prioritization of sub-groups (age, comorbidities, or prioritizing essential workers). For example, Israel was an especially early adopter of the vaccine and prioritized older ages and health risks (5,6). Two months after the initiation of the first vaccination campaign, almost 69% of the individuals eligible for vaccination (age > 18 years), and 81% of individuals older than 60 years, were fully vaccinated (received two doses) in Israel (6).

New variants of the SARS-CoV-2 are constantly evolving worldwide (7,8); some have become the locally or globally predominant variants. For example, in April 2021, the Delta variant emerged from India and quickly became the predominant variant in many parts of the world (9). In Israel, which was among the first countries to reach high vaccination coverage by the end of March 2021, vaccine breakthroughs by the Delta variant were already common during July 2021. Analyses of the breakthrough data pointed to the importance of waning vaccine immunity (10). As a measure to mitigate the Delta variant outbreak while minimizing lockdowns, Israel administered a third dose of the COVID-19 vaccine. Statistical and mathematical models have demonstrated that the booster campaign was instrumental in controlling the Delta resurgence (11–13). However, to estimate the efficiency of other, untested vaccination strategies in conjunction with NPIs, realistic simulation tools are needed.

Model-based analyses have been used to help assess the effectiveness of different NPIs and vaccination strategies (14,15), as controlled experiments are impossible and retrospective data analysis is often lacking. Most modeling studies of SARS-CoV-2 employ compartmental models, such as the differential equation-based SEIR (Susceptible - Exposed - Infected - Recovered/ Removed) model. These models categorize all individuals into a few compartments, not considering the variance in their roles in the outbreak.

An alternative, complementary modeling strategy is individual-based models (IBMs; also called agent-based models, ABMs) with stochastic, interacting autonomous agents representing individuals in the population. IBMs can simulate complex dynamics, the system’s temporal evolution, and generate intricate patterns of behaviors produced by individuals’ interactions. Incorporating fundamental population-specific features can provide insight into systems with small susceptible populations, highly affected by stochasticity (16). This comes at the cost of intense computational demands and decreased interpretability of some results, due to the models’ complexity. However, IBMs have been previously used to simulate the spread of infectious diseases (17–20) and specifically to study the effects of NPIs and vaccination strategies against COVID-19 (21–26).

Here, we developed an IBM populated with COVID-19 parameters. The model simulates outbreaks while considering age-dependent parameters, such as contact matrices, the probabilities of symptomatic and severe disease, and households’ age distribution. Demographic characteristics of a population are especially influential in COVID-19 outbreaks (27,28). Hence, our simulation utilizes the demographic compositions of two Israeli cities with similar population sizes but different household sizes and age distributions, as case studies. First, in the city of Holon, in which about a quarter of the population is <18 years, and the mean household size is approximately 2.8 individuals. In contrast, the city of Bnei Brak, has a much younger and denser population, with approximately half of its population <18 years and average household size of 4.5 individuals. Using these two disparate demographics, we investigate the effects of different vaccination strategies, in combinations with different NPIs, on infections and hospitalizations.

## Methods

### IBM architecture

The IBM is based on three components:

1. <u>Demographics:</u> Each individual in the simulated synthetic population is assigned a certain age, neighborhood, and household. Each individual also has weekly routines (a workplace, school, and time they spend in their neighborhood), determining contacts with other individuals with overlapping routines. Based on the demographic data of either of the two cities in Israel, a synthetic population comprising household members’ joint distribution, ages, employment, and education is generated.
2. <u>Infection dynamics:</u> Daily contact between individuals in the described social setting is simulated (29). Age-heterogeneous contact matrices were used as the risk of exposure to an infected person varies between age groups and locations, representing the probability of contacting persons of another age group. This simulation uses the POLYMOD contact matrix (29). POLYMOD was generated in 2017 and includes contact matrices based on data from 152 countries, including Israel.
3. <u>Disease states (figure 1):</u> At any given time-point, each individual is classified into one of the diseases states: susceptible, infected at a latent state, asymptomatic, presymptomatic, symptomatic, recovered, hospitalized, and deceased (figure S1). Each susceptible individual may change to a latently infected state when contacting an infectious individual. The contact probability, infectiousness probability, and duration distribution in each state are based on previous studies (24). Other parameters, such as susceptibility to infection on contact, relative infectiousness of subclinical cases, and the proportion of hospitalized patients requiring hospitalized care, are based on empirical data and plausible ranges from relevant literature (Table S1).

**Figure 1:**
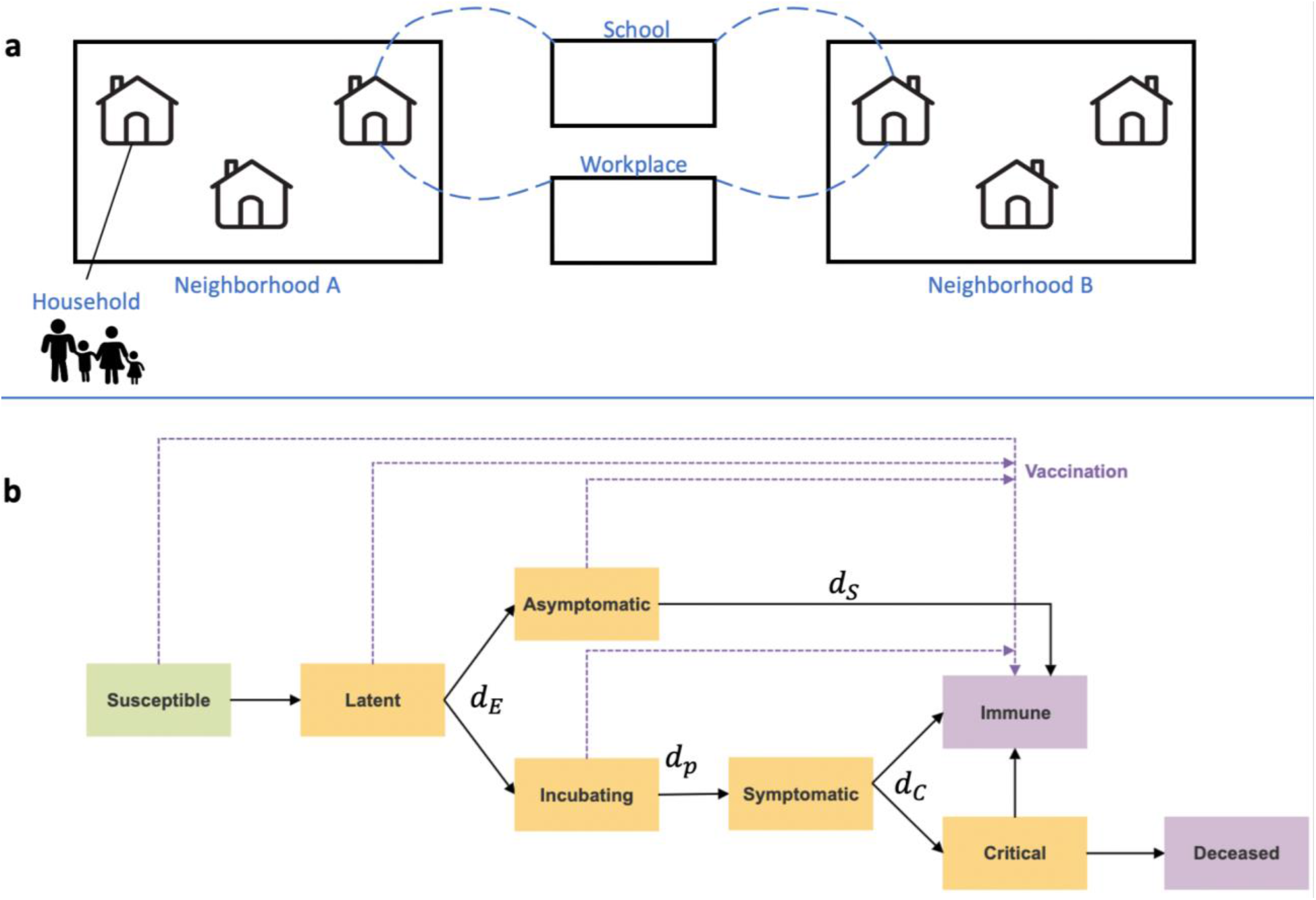
Illustration of the IBM framework. (a) A synthetic population was generated based on Israeli demographics. The population is divided into a hierarchy of social circles, including households and neighborhoods. Contacts between different age groups and settings, such as within their household, school, or workplace, are drawn from data-informed contact matrices (29). (b) Disease progression and immunity states are imposed on each individual in the population. Orange rectangles represent exposed and infectious states and purple rectangles represent removed states. Transition rates between the states are denoted with their parameters, which are given in Table 1.

### Simulation parameters

The main simulation parameters are shown in table 1. Additional model parameters and specifications are given in the supplementary material (table S1).

**Table 1:** Relevant parameters and estimated distribution. The gamma distribution parameterization corresponds to (mean, shape).

| Parameter | Description | Value/<br>Distribution | Reference |
| --- | --- | --- | --- |
| $d_E$ | Latent period (E to I<sub>P</sub>/I<sub>s</sub>; in days) | $\sim gamma(4, 4)$ | (24,30–32) |
| $d_P$ | Duration of preclinical infectiousness | $\sim gamma(1.5, 4)$ | (24,33) |
| $d_C$ | Duration of clinical infectiousness | $\sim gamma(3.5, 4)$ | (24,30–32) |
| $d_S$ | Duration of subclinical infectiousness | $\sim gamma(5, 4)$ | (24,27) |
| | Delay from onset to hospitalization (days) | $\sim gamma(7, 7)$ | (24,34,35) |
| $R_0$ | The basic reproductive number | 3 (2.5, 3.5 as sensitivity analyses) | Calculated from the base infectiousness (36) |
| $u$ | Probability of infection per contact | Calculated from $R_0$ | See details in the SI - “Calibration of $R_0$” section |
| $y_i$ | Probability of clinical symptoms for an infected individual from age group $i$ | Shown in table S1 | |
| $f$ | Relative infectiousness of subclinical cases | 50% | As assumed as in (24,27) |
| $C_{i,j}$ | Number of age-$j$ individuals contacted by an age-$i$ individual (per day) | POLYMOD contact matrix | (29) |
| $Ve$ | Vaccination efficacy against infection and hospitalizations | 90% | (37–39) |
|  | Sensitivity of *Asymptomatic Detection* | 70% | (40) |
|  | Reduction in contacts due to social distancing | Household contacts increase by 25%, Workplace contacts reduce by 25% and within neighborhood contacts reduce by 25%. | As assumed as in (41) |

### Vaccination strategies

The country-wide daily vaccination rate in Israel from the initial vaccination campaign was used to define the daily vaccination rollout for the two evaluated cities, scaled to the size of the cities modeled (4). This led to a daily rollout of 700 vaccines in each city. We deliberately did not use the actual rate of vaccination of each city during the epidemic, so we can directly estimate the effects of demographics, unconfounded by other factors. Individuals that were susceptible, asymptomatic, latent, or incubating post-latent at the time of vaccination were eligible for vaccination. However, individuals that recovered from an asymptomatic infection were not vaccinated. Although this might not reflect reality under policies not testing for antibodies before vaccination, the effect is minor: most asymptomatic cases are below the age of 18 and are not in the eligible age group for vaccination in this simulation. In this simulation framework, individuals are immediately immunized after receiving the vaccine, not reflecting the actual time until immunity, approximately 1-2 weeks (42,43). Thus, a correction for the delayed immunity was implemented by shifting the vaccination timeline by a week (i.e., individuals scheduled to be vaccinated at time *t* were actually vaccinated at time *t+7*).

Four different vaccination strategies were evaluated, keeping the same number of vaccinated individuals per day. Vaccine efficacy (VE) of 90% in preventing infections and hospitalizations was used, as was suggested during the first vaccination campaign (37–39). VE was modeled by limiting the proportion of vaccinated by the value of VE. For example, if VE was set to 90%, then 90% of the individuals originally chosen for vaccination were instead vaccinated daily. The following vaccination strategies were the focus of this study:

1. *General* strategy - vaccinate individuals older than the minimum vaccination age (set at 18) and within the currently vaccinated age group (with random vaccine allocation within the age groups). Meaning that each day, N randomly chosen individuals who meet the criteria for vaccination are being vaccinated.
2. *Neighborhood* strategy - each day, the neighborhood (defined as housing approximately 3,000 individuals) in which the estimated reproductive number (R_t_, using the instantaneous R; for the calculation, see equation S1) is the highest is vaccinated first. Since R_t_ can be calculated only after the first infections are recovered and secondary infections are present, during the first vaccination period, the *Neighborhood* strategy vaccinates from the oldest age group and descends until enough data to calculate the R_t_ is available.

Each strategy was tested with three types of age prioritization: a descending order, where the oldest age group is vaccinated first; an ascending order, where the youngest age group is vaccinated first; and no age prioritization (i.e., vaccination was done randomly in proportion to the age distribution). Results for the latter prioritization strategy were left for the supplementary material (figure S4). Other, less successful, strategies were also implemented and examined, and their details and results are presented in the supplementary material under “vaccination strategies”. These included: a *Household* strategy, wherein individuals in a prioritized age group are vaccinated by clusters of households; an *All At Once* strategy, in which an entire household is vaccinated if it contains at least one individual in the currently prioritized age group.

### NPI modeling

The NPIs in this simulation are modeled by lowering the contact rates between the relevant groups. For example, school closure is modeled by lowering connections between school-aged individuals, and house quarantine is modeled by reducing contacts with the general public while increasing contacts within the household. Compliance with the NPIs is modeled by introducing a parameter governing the proportion of individuals complying with each NPI.

Several NPIs have been investigated. The two NPIs yielding reduction in both infections and hospitalizations without forcing a complete lockdown were chosen to be investigated in combination with the vaccination strategies. In both NPIs, social distancing was implemented in the population. Other interventions were implemented only for specific individuals:

1. *Household isolation* also includes the isolation of symptomatic cases and their household members.
2. *Asymptomatic Detection* includes isolation of symptomatic cases and their household members, and in addition, test and isolation of asymptomatic cases below the age of 18. To consider compliance and the tests’ imperfect sensitivity, only 70% of asymptomatic cases in children in individuals < 18 were detected and isolated.

### Simulation configurations

The main simulation configurations examined and presented in the main text were:

- City demographics (Holon, Bnei Brak)
- Vaccination Strategies
  - Strategy (*General, Neighborhood*)
  - Age prioritization (youngest to oldest, oldest to youngest, random)
- NPI (*Household Isolation, Asymptomatic Detection*)

### Evaluation metrics

Two metrics were calculated for each scenario to compare the performance of the different vaccination strategies: the total number of infections and the total number of hospitalized cases per 100k individuals, averaged over 500 simulations. These metrics were chosen to reflect disease spread and morbidity, and hence reflect public health and economic costs. All simulations were run for 150 days to emulate simplified SARS-CoV-2 variant outbreak dynamics. Additionally, this period corresponds to the eligibility criterion of the minimal time elapsed from vaccination to a booster shot, as was set in Israel (10).

The mean differences in the total infections and hospitalizations, across simulation repetitions, were compared for pairs of vaccination strategies. The central limit theorem was used to derive 95% confidence intervals (CI) for these comparisons, assuming unequal variances.

## Results

First, typical outbreak dynamics under the *Asymptomatic Detection* intervention are shown in figure 2. The results are presented for both demographics, and include the number of infected (a,b) and hospitalized (c,d) individuals, and the reproductive number R_t_ (e,f), over time. An analogous figure for the *Household Isolation* intervention can be found in the SI figure S8.

**Figure 2:**
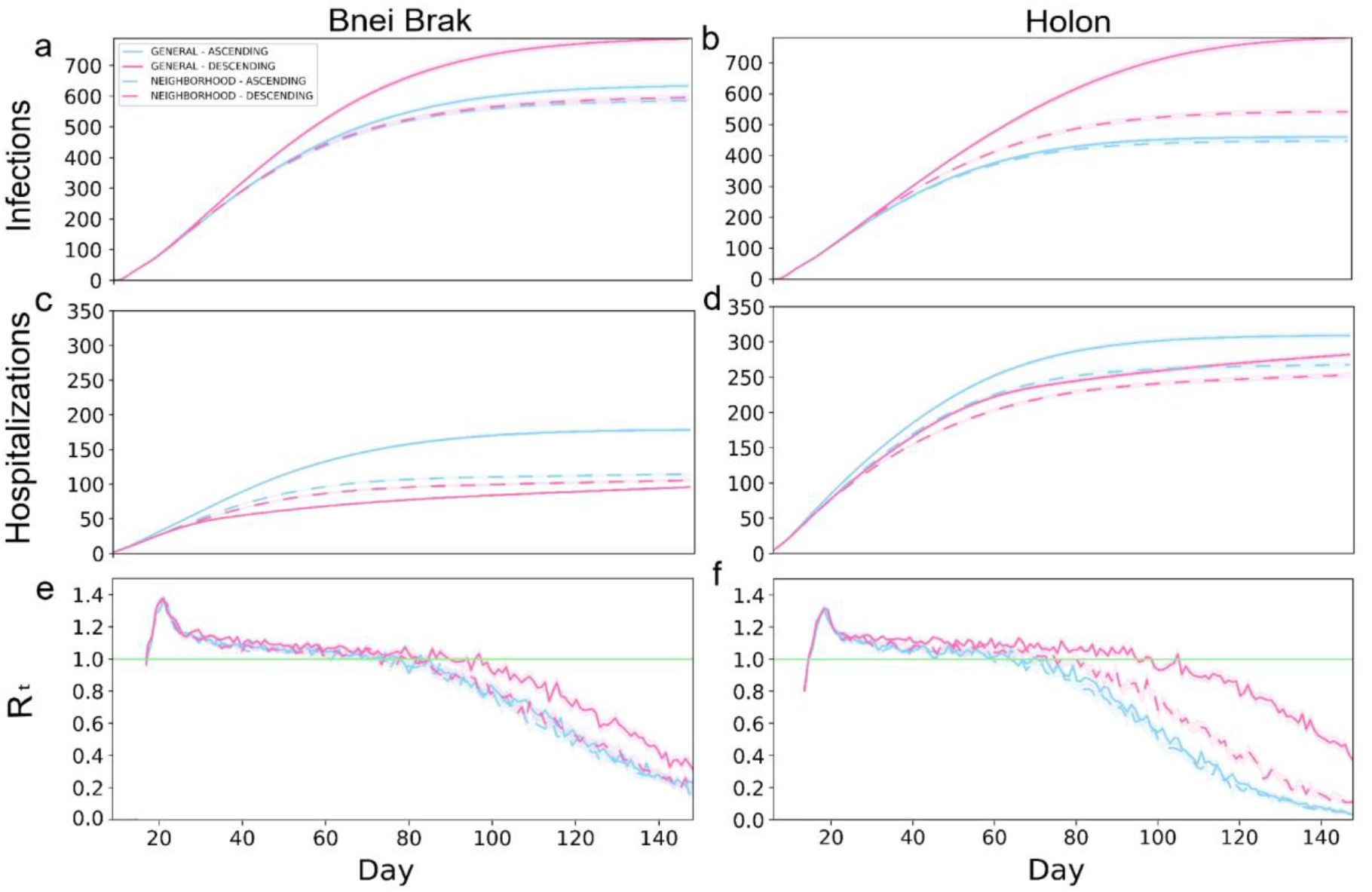
Typical outbreak dynamics under different vaccination strategies and demographic structures. The cumulative number of new infections per 100k (a,b), hospitalizations per 100k (c,d), and R_t_ (e,f), are shown. The left- and right-hand columns present the results under the demography of Bnei Brak and Holon, respectively. Each panel presents the daily mean of 500 simulations, and the shaded regions around the curves represent the standard error of the mean.

The two *Neighborhood* vaccination strategies, prioritizing neighborhoods with high transmission rates, yielded low infection rates throughout the simulations. The *Neighborhood Descending* strategy allowed for low hospitalization rates, while keeping the total infections lower than the *General* strategies and similar to the *Neighborhood Ascending* strategy. Furthermore, while the *General Ascending* strategy resulted in only a few more infections than both *Neighborhood* strategies, it resulted in almost twice as many hospitalizations.

Simulating Holon’s demographics, the *Neighborhood Descending* strategy resulted in the best trade-off between both criteria compared to the other combinations (figure 2 b,d). While this strategy did not result in the lowest number of infections, it yielded relatively low numbers in both criteria. On the other hand, the *General Descending* strategy resulted in almost twice as many infections as the two ascending strategies. Moreover, this strategy resulted in about 1.6-fold infections as its analogous *Neighborhood* strategy. When simulating the city demographics of Bnei Brak, the *Neighborhood Descending* strategy again resulted in an advantage over its analogous *General* strategy, but with a less substantial difference than in the demographics Holon. Thus, the *Neighborhood Descending* strategy yielded a reasonable trade-off between the two metrics (total infections and total hospitalizations), for both cities.

To complete the picture, we also examined the daily reproductive number (compare figure 2 e to f). Implementing *Asymptomatic Detection* with any vaccination strategy resulted in a decline in the reproductive number in both cities, with a faster decline in Holon, eventually dropping below 1 (i.e., the number of infections is reducing). In contrast, the R_t_ values under the *Household Isolation* intervention did not drop below 1 until the completion of the simulation (figures S5, S6). Lastly, while comparing the overall performance of the different strategies, it appears that there is larger variability between the strategies in the total infections and the R_t_-values in Holon (compare a to b, and e to f in figure 2).

In figure 3, we present a comparison of the distributions of the total infections and hospitalizations when applying the *Asymptomatic Detection* and the *Household Isolation* NPIs, and for different vaccination strategies. All comparisons were performed using the number of cases per 100,000 individuals. Compared to *Household Isolation, Asymptomatic Detection* limited both epidemiological metrics assessed: it resulted in approximately 1/8 of the total infections in Bnei Brak’s demographics, less than half the infections in Holon’s demographics, and less than 1/2 of the hospitalized cases in both demographics.

**Figure 3:**
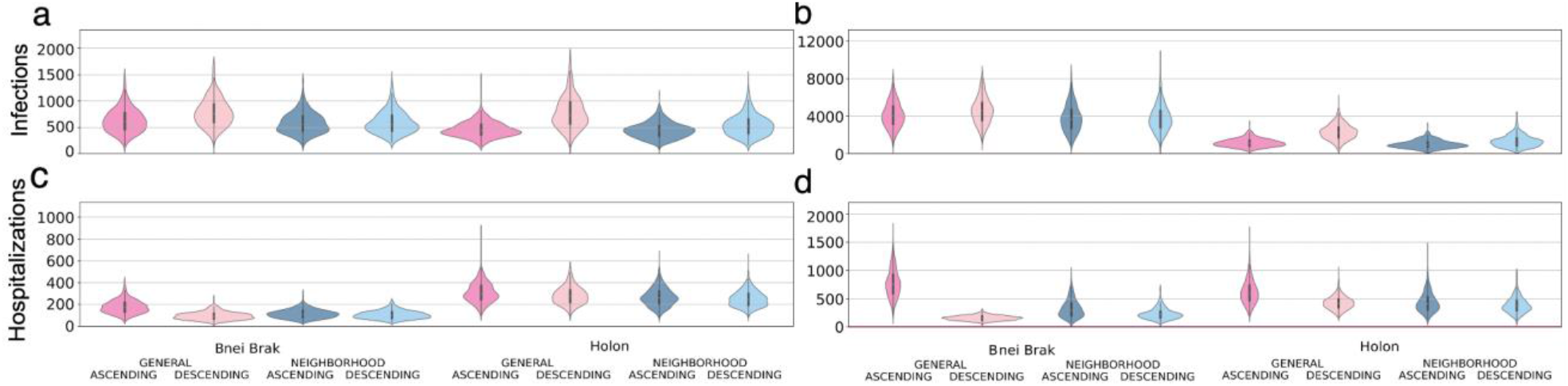
Examining the efficiency of the two main NPIs. Each panel shows violin plots of the number of infected (a and b) or hospitalized (c and d) per 100k individuals at the end of 500 simulations. The plots are further stratified by the application of the *Asymptomatic Detection* (a and c) or *Household Isolation* (b and d) interventions.

Simulating Bnei Brak’s demographics, under the *Asymptomatic Detection* intervention, both *Neighborhood* strategies led to fewer infections per 100,000 individuals compared to the *General Ascending* strategy (−48.1, 95% CI [-74.3, -21.9] for *Neighborhood Ascending*, and -37.6, 95% CI [-64.5, -10.7] for *Neighborhood Descending*). Furthermore, the *Neighborhood Descending* strategy also resulted in fewer infections \compared to the *General Descending* strategy (−191.8, 95% CI [-221.3, -162.3]). The number of infections under *Neighborhood Ascending* and *Neighborhood Descending* were very similar and not statistically different (−10.5, 95% CI [-35.6, 14.6]). Comparing the total hospitalizations, rates were higher for the *Neighborhood Descending* strategy than for the *General Descending* strategy (9.8, 95% CI [5.3, 14.3]). Still, the *Neighborhood Descending* strategy performed better than the *Neighborhood Ascending* (−8.8, 95% CI [-13.6, -4.0]) and the *General Ascending* strategies (−72.6, 95% CI [-78.8, -66.5]). Moreover, the *Neighborhood Ascending* strategy led to fewer hospitalizations than the *General Ascending* strategy (−63.9, 95% CI [-70.5, -57.5]).

We observed a similar trend when applying the *Household Isolation* intervention: *Neighborhood Ascending* reduced infections better than the *General Ascending* (−334.0, 95% CI [-508.8, - 167.1]) and *General Descending (*-800.5, 95% CI [-975.3, -625.6]) strategies. Similar to the results of applying *Asymptomatic Detection*, the *Neighborhood Descending* strategy reduced infections better than both the *General Ascending* (−328.6, 95% CI [-498.4, -158.8]) and *General Descending* (−791.1, 95% CI [-964.9, -617.4]) strategies. The difference between the *Neighborhood Ascending* and the *Neighborhood Descending* strategies was again not significant (−9.3, 95% CI [-185.6, 167.0]). When examining the ability to limit total hospitalizations, both *Descending* strategies were better than their parallel *Ascending* strategies (−93.4, 95% CI [-109.5, -77.3] for the difference between the *Neighborhood Descending* and *Neighborhood Ascending* strategies and -619.25, 95% CI [-641.1, -597.4] for the difference between the *General* strategies). While comparing both *Descending* strategies, the *Neighborhood Descending* strategy resulted in more hospitalization than the *General Descending* strategy (73.6, 95% CI [64.5, 82.8]).

When simulating the city demographics of Holon, there were no significant differences in infection rates between the *Neighborhood Ascending* and the *General Ascending* strategies (−13.3, 95% CI [-31.3, 4.6]). Still, both *Neighborhood* strategies performed better than the commonly used *General Descending* strategy (−333.9, 95% CI [-363.2, -304.5] for *Neighborhood Ascending* Vs. *General Descending*; and -238.9, 95% CI [-270.6, -207.2] for *Neighborhood Descending* Vs. *General Descending*). *Neighborhood Descending* performed better in reducing hospitalizations than all other strategies (−56.0, 95% CI [-66.2, -45.8] for *Neighborhood Descending* Vs. *General Ascending;* -29.2, 95% CI [-38.6, -19.7] for *Neighborhood Descending* Vs. *General Descending*; and -14.7, 95% CI [-24.4, -5.0] for *Neighborhood Descending* Vs. *Neighborhood Ascending*).

Applying *Household Isolation* to the city demographics of Holon, the *Neighborhood Ascending* strategy resulted in fewer infections than its analogous *General Ascending strategy (*-154.7, 95% CI [-207.2, -102.3]) and an even more substantial reduction compared to the *General Descending* strategy (−1308.5, 95% CI [-1388.0, -1229.1]). Comparing the total hospitalizations, the *Neighborhood Descending* strategy had an advantage compared to all other strategies (−234.8, 95% CI [-256.3, -213.2] for *Neighborhood Descending* Vs. *General Ascending*; -41.1, 95% CI [- 56.5, -25.7] for *Neighborhood Descending* Vs. *General Descending*; and -52.8, 95% CI [-71.9, - 33.7] for *Neighborhood Descending* Vs. *Neighborhood Ascending*).

We further sought to examine whether vaccination strategies should be uniformly or differentially applied to locations with differing demographics. To this end, we independently simulated outbreaks under both demographic compositions (i.e. no spillover between the two populations) and both NPIs and summed the infections and hospitalizations per 100,000 people. These results are presented in figure 4, under either uniform vaccination strategies, where the same strategy is applied in both cities, or under a different strategy for each demographic composition. When applying *Household Isolation*, the optimal strategy in terms of the combined number of hospitalizations was *General Descending* under the Bnei Brak demography and *Neighborhood Descending* under the Holon demography. Similarly, when applying the *Asymptomatic Detection* intervention, using the same combination of strategies resulted in the lowest combined number of hospitalizations.

**Figure 4:**
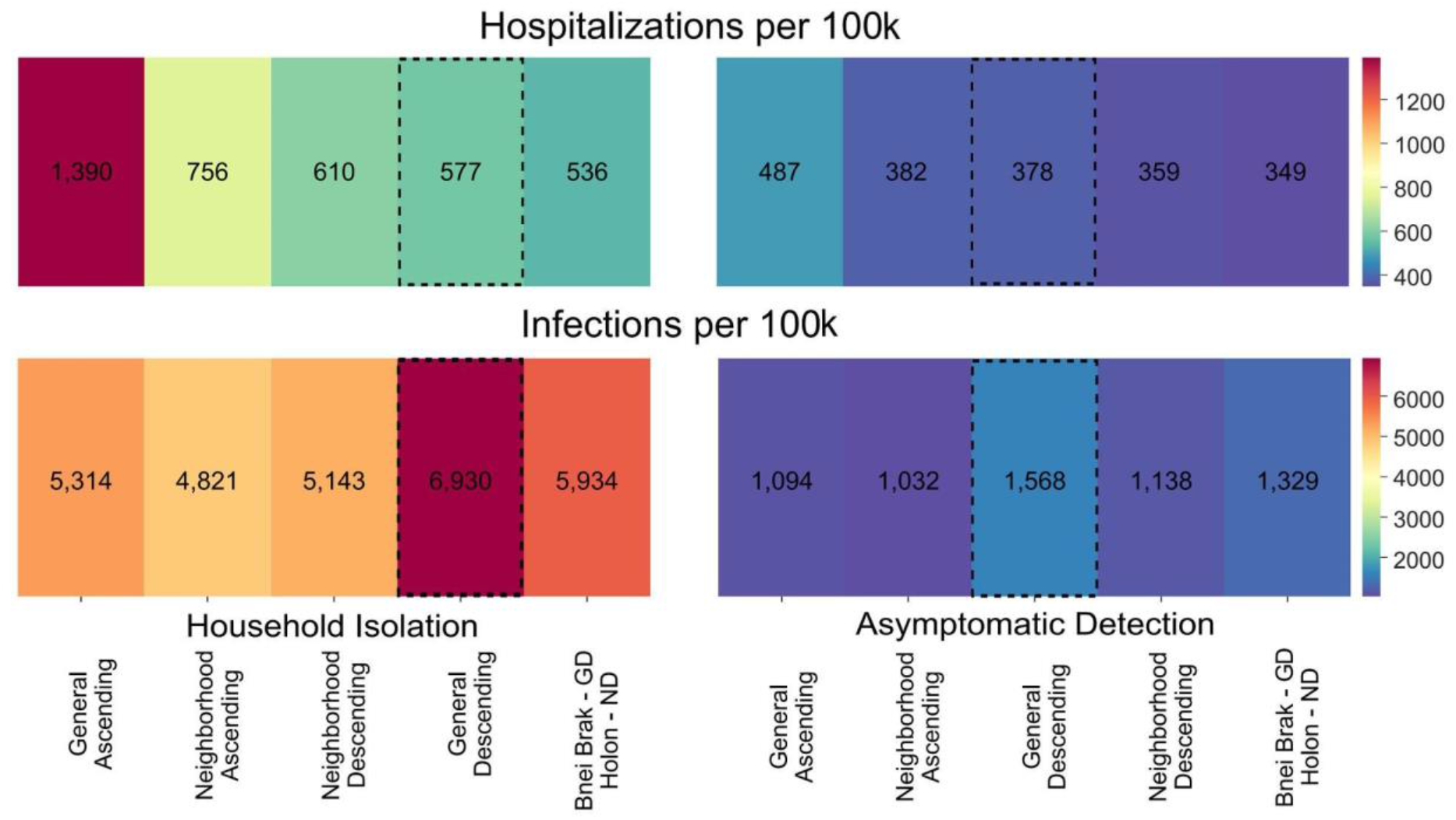
Applying vaccination strategies uniformly and differentially between demographics. Colors represent the mean hospitalizations (top) and infections (bottom) per 100k individuals, combined for simulations under both the Holon and Bnei Brak demographic compositions. Results are shown using either the same vaccination strategy in both demographic compositions or the best strategy for each demographic composition, under *Household Isolation* (left) or *Asymptomatic Detection* (right). The numbers represent the mean of 500 simulations per scenario. The vaccination strategies are sorted according to hospitalizations. The *General Descending* strategy is highlighted as it is the most commonly used strategy in most countries. ND - *Neighborhood Descending*, NA - *Neighborhood Ascending*, and GD - *General Descending*.

## Discussion

This study employs a realistic individual-based model of COVID-19 dynamics to investigate essential epidemic outcomes under different vaccination strategies. The unique structure of our IBM enabled us to compare complex vaccination strategies that target subpopulations and their interplay with applications of different NPIs. These investigations would have been extremely difficult to implement using classic SEIR-based compartmental models. Moreover, we explicitly considered differing demographic structures, incorporating population structures from two demographically distinct Israeli cities.

Our results are consistent with the widely accepted notion that prioritizing the elderly for vaccination reduces hospitalizations under a broad range of conditions. Indeed, this has been demonstrated in theoretical studies (15) and was the strategy of choice in many countries (44), following the finding that older individuals have higher chances of developing severe COVID-19 given infection. On the other hand, we showed that prioritizing vaccination for younger individuals reduced infections. This is due to the high rate of social interactions of younger individuals, coupled with their higher rates of asymptomatic infections (45,46), leading to lower probabilities of case isolations.

We further examined the effects of NPIs and vaccination strategies on the two distinct demographic compositions. We found that the same interventions and vaccination strategies produced different results in the selected population structures. Under the ‘older’ demography of Holon, the different vaccination strategies mainly affected the number of infections and, correspondingly, the *R*_*t*_ values. With respect to NPIs, actively detecting asymptomatic infections (*Asymptomatic Detection*) led to a significantly lower number of infections and hospitalizations than isolating household members of symptomatic cases (*Household Isolation*).

Several reasons could explain the success of the *Asymptomatic Detection* intervention. First, younger individuals tend to be more mobile and have more contacts per day (47), leading to higher chances of secondary infections for each asymptomatic case. Second, since younger individuals are less prone to present symptoms (27,48), their abundance may reduce the effectiveness of NPIs that mainly target symptomatic cases (49). Altogether, actively testing to detect asymptomatic infections can be valuable in demographic settings with a younger population.

Nevertheless, applying the *Asymptomatic Detection* intervention is not trivial. First, it requires compliance from parents to test their children frequently. Second, it is expensive to implement due to the high use of testing kits. On the other hand, the *Asymptomatic Detection* intervention successfully reduced both the number of infections and hospitalizations compared to *Household Isolation*. For example, under the demographics of Bnei Brak, the *Household Isolation* intervention resulted in an eight-fold increase in infections and almost a two-fold increase in hospitalizations relative to applying *Asymptomatic Detection*. Accordingly, when aiming to limit the number of hospitalizations and reduce the burden on intensive care units, *Asymptomatic Detection* should be investigated as a useful NPI, especially in settings with a young population structure.

Under both demographic settings, the *Neighborhood* strategies were less affected by the order of vaccination than the *General* strategies. A possible explanation is that the *Neighborhood* strategies focused all of the vaccines on a single neighborhood at a time. Thus, it allowed for a more rapid shift from the prioritized age group to other age groups within the neighborhood. Moreover, for the demographics of Holon, the *Neighborhood Descending* strategy performed better than any other strategy, under both NPIs, to reduce hospitalizations and better than the *General Descending* strategy in reducing infections. Under the demographics of Bnei Brak, the *Neighborhood Descending* strategy performed best in reducing infections and resulted in the second-lowest total hospitalizations. Notably, the *General Descending* strategy is the most used vaccination strategy worldwide (44). However, while reducing hospitalizations, the *General Descending* strategy performed the worst at reducing infections compared to the other vaccination strategies simulated. We hence propose that using the *Neighborhood Descending* strategy as a potential alternative can provide a good trade-off between total hospitalizations and infections.

The *Neighborhood* strategies are analogous to “targeted geographic vaccination”. This previously-used vaccination strategy focuses on areas, neighborhoods, or villages with higher infection rates (50) and was successfully used in the Ebola outbreak in Chow in South Kivu (51). Moreover, it was shown that the targeted geographic vaccination was more effective than both mass and ring vaccination (52). Hence, focusing on a larger population unit than a primary circle of contacts, such as a neighborhood, has empirical support of potential efficiency - aligned with our results. The disadvantages of the *Neighborhood* vaccination strategy are its logistical demands and public perception. The selected neighborhood and prioritization within the neighborhood can change rapidly, requiring efficient information flow and high compliance from the neighborhoods’ residents. To overcome the logistical difficulties, small neighborhoods can be combined into larger clusters based on their *R*_*t*_. Furthermore, the selection of the next cluster to vaccinate can be made less frequently (e.g., every week). On the other hand, targeting specific subpopulations before others can be perceived negatively by the public. Subpopulations offered the vaccine early on might hesitate to comply for fear of side effects (53); alternatively, sup- populations vaccinated later on may feel discriminated against (54).

Finally, we sought to understand whether applying mixed strategies to different populations could be beneficial. While a uniform vaccination strategy is usually chosen across entire countries, we showed a potential advantage in combining different strategies for different sub-populations. For example, while applying the *Household Isolation* intervention, using a combined strategy resulted in 7% fewer hospitalizations compared to the best uniform strategy, which in this case was the commonly used *General Descending* strategy. This demonstrates the potential benefit of selecting the optimal strategy per city, county, or other lower-level localities, as countries are not uniform in their demographics. However, similar difficulties to those elaborated above regarding the *Neighborhood* strategies apply when implementing combinations of vaccination strategies in different localities.

We also note that our assumed form of VE imposes potential limitations on the generalizability of our study. Since we model vaccination by moving individuals directly to the recovered compartment, our model refers to vaccination as equally efficient against infections and developing severe disease given infection. Consequently, our study is mainly relevant for scenarios where VE is similar with respect to both of these outcomes. Indeed, this assumption held in the initial vaccination campaigns against the first SARS-CoV-2 variants (37–39). Future work should focus on expanding this model and decomposing VE so it can assume different values against infection and severe disease.

To conclude, we have shown that subpopulations with different demographic compositions may require different vaccination strategies. Our study emphasizes the importance of tailoring a strategy to the unique characteristics of a subpopulation’s demographics (e.g., household size distribution and age distribution) instead of following a uniformly applied strategy. We also demonstrated the effectiveness of vaccination strategies targeting highly infected localities and of NPIs that actively seek asymptomatic infections. The presented simulation framework and our findings can help better design future responses against emerging SARS-CoV-2 variants and other pathogens.

## Data Availability

The datasets presented in this study can be found in online repositories. The names of the repository/repositories and accession number(s) can be found in the article/supplementary material.

https://github.com/TAU-COVID19/coderona-virus

## Acknowledgments

We thank Ronen Olinky, Tom Kalavari, and the rest of the scenarios team recruited for the early COVID-19 response by the Gertner Institute for Epidemiology and Health Policy Research, for helping with the design and coding of an early version of the model.

## Data and code availability

All used data are publicly available or can be found, together with the code used for the simulations at https://github.com/TAU-COVID19/coderona-virus.

