## Supplementary Information for "Assessing COVID-19 vaccination strategies in varied demographics using an individual-based model"

### Supplementary material

#### SEIR model, parameters, and their distribution

The SEIR model compartments were implemented with modifications. Our model has the following eight compartments (figure 1 in the main text):

1. Susceptible - In the beginning, all individuals are susceptible to the disease, except for a chosen number of random individuals that are infected.
2. Latent/Exposed - Susceptible individuals move to the latent compartment after encountering an infectious individual.
3. Infected individuals are either:
    a. Asymptomatic.
    b. Incubating post latent and then Symptomatic.
4. Critical - Some cases are hospitalized and move to a critical stage.
5. Immune - including:
    a. Recovered from a disease.
    b. Vaccinated.
    c. Deceased.

It was assumed that all recovered individuals were fully immuned and could not be reinfected. It was also assumed that asymptomatic and latent stages are half as infectious as the symptomatic and hospitalized stages. Vaccination was implemented by moving a fraction of individuals directly to the immune compartment each day. Only individuals with no indication of being sick can be vaccinated. In other words, people in susceptible, latent, incubating post latent, or asymptomatic states can be vaccinated. In our simulation framework, even recovered cases from an asymptomatic infection were not vaccinated, although, in reality, there would probably not be a way to separate them from the susceptibles.

The probability of symptomatic disease given an infection per age group was calculated using a linear interpolation of the estimates by Ma, Qiuyue, et al. (1), to account for a higher resolution of the age groups used in our model.
Table S1 presents the probability of infected individuals of different age groups to present clinical symptoms.

| Age group | 0-10 | 10-20 | 20-30 | 30-40 | 40-50 | 50-60 | 60-70 | 70-80 | 80+ |
|---|---|---|---|---|---|---|---|---|---|
| *Probability of symptoms* | 0.35 | 0.42 | 0.49 | 0.56 | 0.63 | 0.7 | 0.76 | 0.84 | 0.87 |

**Table S1: Probability of clinical symptoms on infection for age group $i$ ($y_i$).**

### Creating a population

Data of two city demographics were incorporated into the model, including age and household size distribution. In particular, the number of individuals in each group was created using a heuristic rejection sampling algorithm based on demographic data from the central bureau of statistics (see “population generation” in the linked code repository).

The generation of households followed town-specific demographic information from the Central Bureau of Statistics. First, lists of the below demographic information were created:

1. Age distribution of the population in the city. The code divides the ages according to the city’s age distribution (https://www.cbs.gov.il/en/subjects/Pages/Population-in-Localities.aspx).
2. The number of houses and the distribution of household sizes (https://www.cbs.gov.il/en/subjects/Pages/Households.aspx).
3. Percentage of households with individuals above the age of 65 (https://www.cbs.gov.il/en/subjects/Pages/Households.aspx).
4. Distribution of the number of children in each household (https://www.cbs.gov.il/en/subjects/Pages/Households.aspx).

Then, households are generated, updating the lists after each household generation (so that the next household will be generated based on the distribution remaining).

### Calibration of $R_0$

The model was calibrated to achieve an $R_0$ value close to 3, as was estimated for the first variant with the initial pandemic data (2). We used our simulation to find the base infectiousness per contact that corresponds to this value of $R_0$. We chose a value that allowed for $R_0$ close to 3 in both cities: 0.09 (figure S1). We also tested our vaccination strategies with lower base infectiousness of 0.07 and 0.11, corresponding to $R_0$ of about 2.5 and 3.5, as a sensitivity analysis.

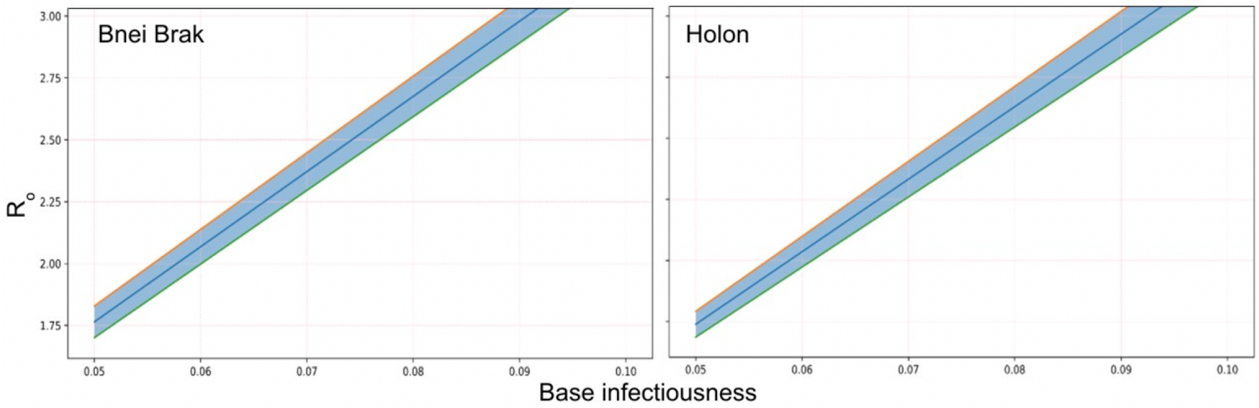


**Figure S1: $R_0$ as a function of the base infectiousness in the cities of Bnei Brak and Holon.** The blue line represents the mean values of the simulation repetitions, and the blue shading represents one standard error from the mean.

#### Calculation of $R_t$

The effective reproductive number is the expected number of new infections caused by one infectious individual at time t. Calculating the effective reproductive number involves right censoring. Hence, the instantaneous $R_t$, which is based on past data, is often used in practice to estimate the expected number of new infections at time t, given that all conditions affecting infection remain the same (3). On the other hand, since the instantaneous $R_t$ "looks backward" and uses the information from prior t days for its calculation, it is not meaningful at the beginning of the simulation. Here we use information from the prior 14 days, as only a minor fraction of the infections are still infectious after this period (4). For this reason, we show $R_t$ data only from the 14th day in figure 2 and figure S8.

The instantaneous reproductive number is defined as the expected number of secondary infections at time t, divided by the number of infected individuals, each scaled by their relative infectiousness at time t. The relative infectiousness is a function of the generation interval and the time passed since the infection. In our model, we use the *Cori et al.* method to estimate $R_t$ from the simulations (5), as was recommended in *Gostic et al.* (3):

$$R_t = \frac{I_t}{\sum_{s=1}^{t} (I_{t-s} \times w_s)} \qquad \text{(S1)}$$

Where $I_t$ is the number of infections on day t and $w_s$ is the relative infectiousness on day t, defined by the generation interval. In other words, this estimator describes the number of new cases on day t, relative to the number ($I_{t-s}$) individuals who became infected s days before t, weighted by the current infectiousness of individuals that were infected *s* days ago.

#### Vaccination strategies

We investigated two additional vaccination strategies to those presented in the main text: *Household* strategy and *All At Once* strategy:

1. *Household* strategy - Aimed to target the households, as this is where individuals spend most of their time. Each day, M households are randomly chosen, and all individuals in the households that belong to the currently prioritized age group are being vaccinated.
2. *All At Once* strategy - Similar to the *Household* strategy, but instead of vaccinating only the current age group, it vaccinates the entire household if at least one household member is in the currently vaccinated age group.

Moreover, we investigated all the strategies without favoring any age group, selecting individuals at random above the minimum age of vaccination.

Figures S2 and S3 show the number of infections and hospitalizations using the *Household* and *All At Once* vaccination strategies. Figure S4 examines the *General* and *Neighborhood* vaccination strategies, while no age group is prioritized.

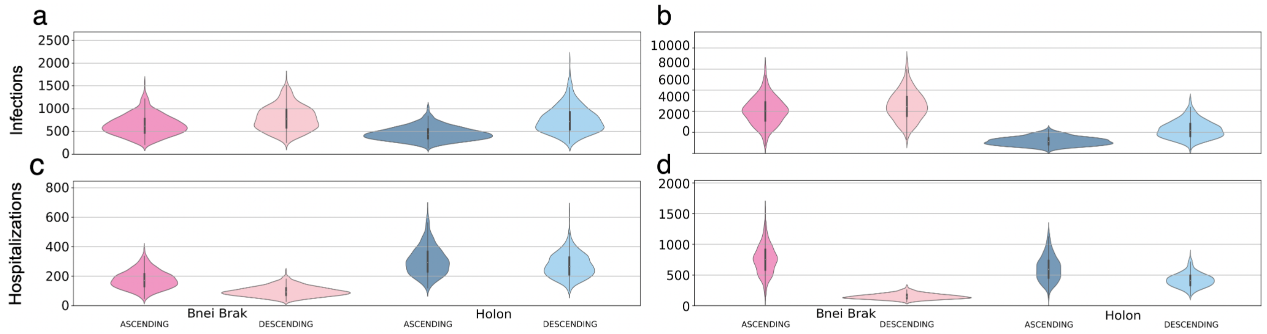


**Figure S2: Examining the efficiency of the *Household* vaccination strategy, together with the two main NPIs.** Each panel shows violin plots of the number of infected (a and b) or hospitalized (c and d) per 100k individuals at the end of 500 simulations. The plots are further stratified by the application of the *Asymptomatic Detection* (a and c) or *Household Isolation* (b and d) interventions.

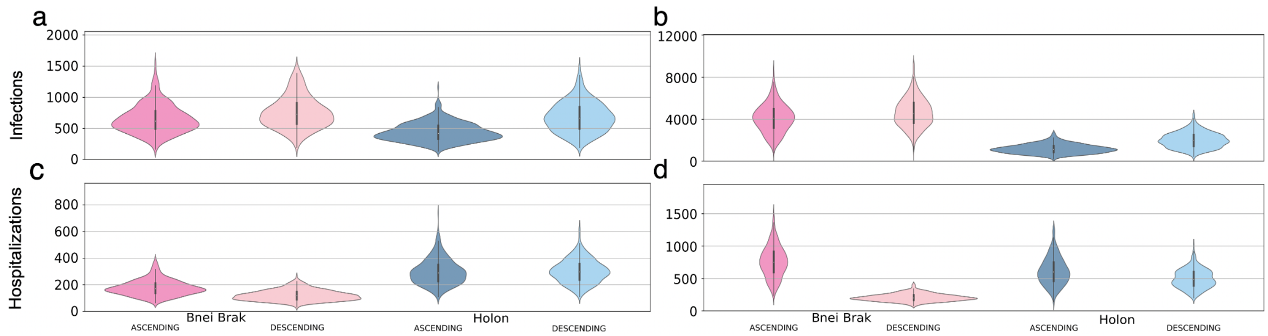


**Figure S3: Examining the efficiency of the *All At Once* vaccination strategy, together with the two main NPIs.** Each panel shows violin plots of the number of infected (a and b) or hospitalized (c and d) per 100k individuals at the end of 500 simulations. The plots are further stratified by the application of the *Asymptomatic Detection* (a and c) or *Household Isolation* (b and d) interventions.

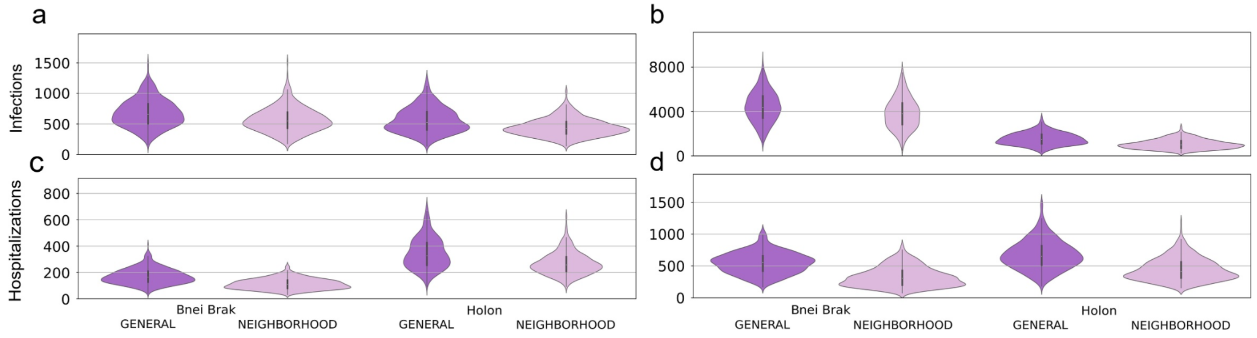


**Figure S4: Examining the efficiency of the two main NPIs without age prioritization.** Each panel shows violin plots of the number of infected (a and b) or hospitalized (c and d) per 100k individuals at the end of 500 simulations. The plots are further stratified by the application of the *Asymptomatic Detection* (a and c) or *Household Isolation* (b and d) interventions.

### Sensitivity analyses

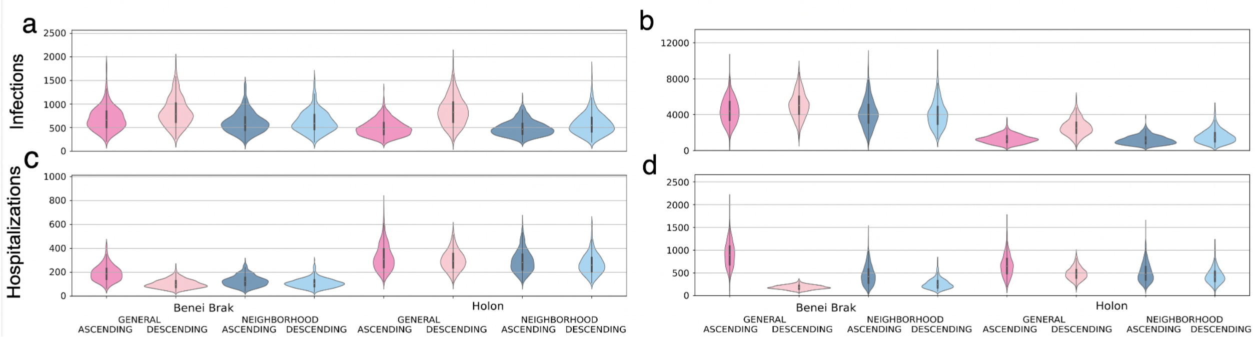


**Figure S5: Examining the two main NPIs under vaccine efficiency of 80%.** Each panel shows violin plots of the number of infected (a and b) or hospitalized (c and d) per 100k individuals at the end of 500 simulations. The plots are further stratified by the application of the *Asymptomatic Detection* (a and c) or *Household Isolation* (b and d) interventions.

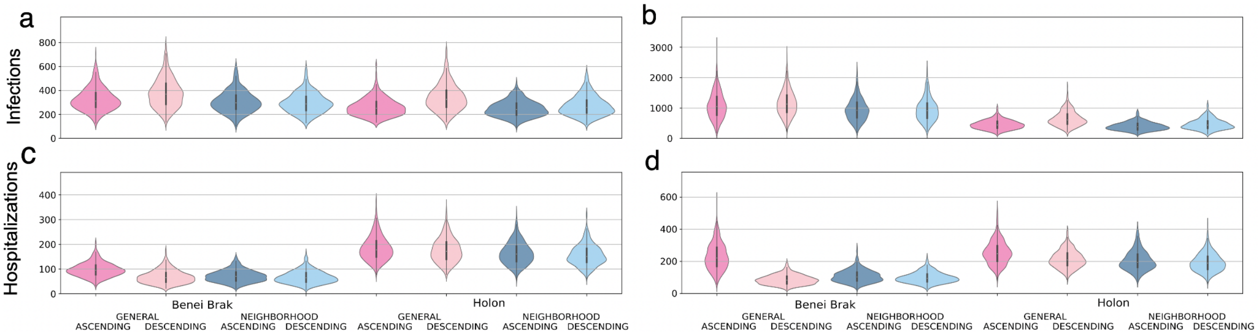


**Figure S6: Examining the two main NPIs under base infectiousness of 0.07 ($R_0 \approx 2.5$).** Each panel shows violin plots of the number of infected (a and b) or hospitalized (c and d) per 100k individuals at the end of 500 simulations. The plots are further stratified by the application of the *Asymptomatic Detection* (a and c) or *Household Isolation* (b and d) interventions.

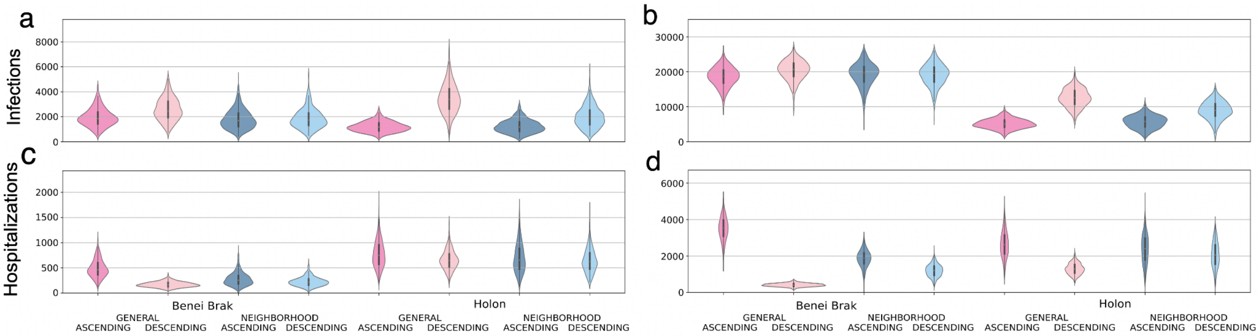


**Figure S7: Examining the two main NPIs under base infectiousness of 0.11 ($R_0 \approx 3.5$).** Each panel shows violin plots of the number of infected (a and b) or hospitalized (c and d) per 100k individuals at the end of 500 simulations. The plots are further stratified by the application of the *Asymptomatic Detection* (a and c) or *Household Isolation* (b and d) interventions.

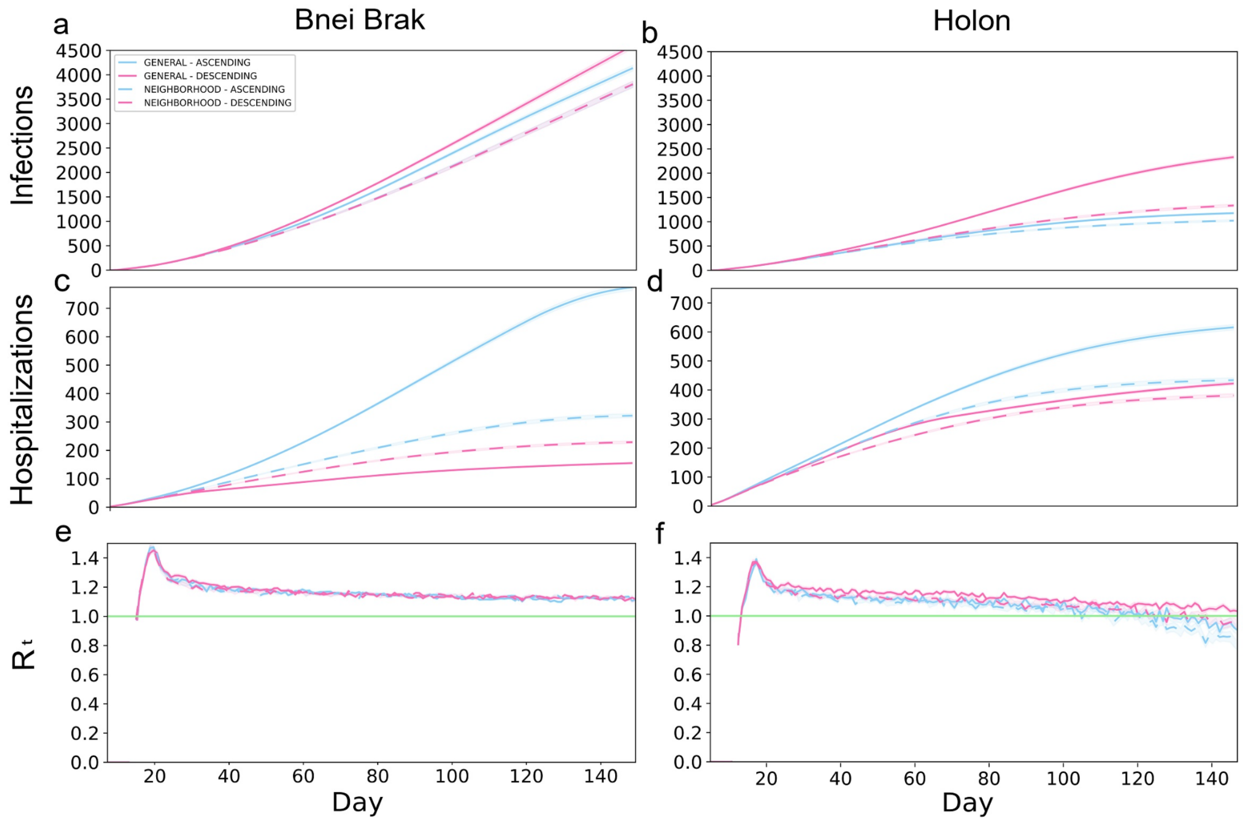


**Figure S8: Typical outbreak dynamics under different vaccination strategies and using *Household Isolation* intervention.** The cumulative number of new infections per 100k (a,b), hospitalizations per 100k (c,d), and $R_t$ (e,f), are shown. The left- and right-hand columns present the results under the demography of Bnei Brak and Holon, respectively. Each panel presents the daily mean of 500 simulations, and the shaded regions around the curves represent the standard error of the mean.
